# Frailty increases the risk of adverse outcomes among 38,950 UK Biobank participants with prediabetes: A prospective cohort study

**DOI:** 10.1101/2022.12.11.22283325

**Authors:** Xingqi Cao, Xueqin Li, Jingyun Zhang, Xiaoyi Sun, Gan Yang, Yining Zhao, Shujuan Li, Emiel O. Hoogendijk, Xiaofeng Wang, Yimin Zhu, Heather Allore, Thomas M. Gill, Zuyun Liu

## Abstract

**Background:** We aimed to systematically evaluate the associations of frailty, a simple health indicator, with risks of multiple adverse outcomes in late life among adults with prediabetes.

**Methods:** We evaluated 38,950 adults aged 40-64 years with prediabetes from the baseline survey of the UK Biobank. Frailty was assessed using the frailty phenotype (FP, 0-5), and participants were grouped into non-frail (FP =0), pre-frail (1≤ FP ≤2), and frail (FP ≥3). Multiple health outcomes were ascertained during a median follow-up of 12 years. Cox proportional hazards regression models were used to estimate the associations.

**Results:** At baseline, 49.1% and 5.9% of adults with prediabetes were identified as pre-frail and frail, respectively. Both pre-frailty and frailty were associated with higher risks of multiple adverse outcomes in adults with prediabetes (P for trend <0.001). For instance, compared with their non-frail counterparts, frail participants with prediabetes had a significantly higher risk (P <0.001) of type 2 diabetes mellitus (T2DM) (hazard ratio [HR]: 1.73), diabetes-related microvascular disease (HR: 1.89), cardiovascular disease (HR: 1.66), chronic kidney disease (HR: 1.76), eye disease (HR: 1.31), dementia (HR: 2.03), depression (HR: 3.01), and all-cause mortality (HR: 1.81) in the multivariable-adjusted models. Furthermore, with each 1-point increase in FP score, the risk of these adverse outcomes increased by 10% to 42%.

**Conclusions:** In UK adults with prediabetes, both pre-frailty and frailty are significantly associated with higher risks of multiple adverse outcomes, including T2DM, diabetes-related diseases, and all-cause mortality. Our findings suggest that frailty assessment should be incorporated into the routine care for middle-aged adults with prediabetes, to improve the allocation of healthcare resources and reduce diabetes-related burdens.

## Introduction

In 2021, the International Diabetes Federation estimated that there were more than 500 million adults with prediabetes among those aged 20-79 years worldwide [1]. As an intermediate hyperglycemia state, prediabetes increases the risk of diabetes [2] and diabetes-related complications [3]; the latter contributes to a large proportion of diabetes-related burden [4, 5]. The 2020 guidelines from the American Diabetes Association (ADA) recommend annual diabetes screening for adults with prediabetes. However, this is challenged by emerging evidence showing the very low rates of diabetes progression among older adults with prediabetes[6]. Conversely, middle-aged adults (i.e., <65 years) with prediabetes should be monitored for adverse outcomes, which is high-value and appropriate [7].

Prediabetes is highly heterogeneous, impeding the application of a one-size-fits-all health management strategy. Recently, a simple health aging indicator — frailty has been demonstrated to be able to predict the risk of adverse outcomes (e.g., cardiovascular disease [CVD] and mortality) [8-11], even in the younger population [12]. Frailty is defined as a state of decreased reserve and resistance to stressors, characterized by the functional decline in multiple systems [8]. Both frailty and disorders of glucose metabolism share common physiological mechanisms, such as insulin resistance [13, 14] and chronic inflammation [14, 15]. A few studies have shown that frailty incidence is slightly higher in prediabetic older adults compared to those with normal glucose metabolism [16]. However, relatively little is known about whether frailty could identify middle-aged adults who are most at risk of adverse outcomes related to prediabetes.

Therefore, we performed a prospective cohort study among 38,950 middle-aged adults with prediabetes from the UK Biobank (UKB). Using a widely validated frailty measurement — frailty phenotype (FP), this study aimed to systematically evaluate the associations of frailty with the risk of multiple adverse outcomes, including incident type 2 diabetes mellitus (T2DM), diabetes-related microvascular disease, CVD, chronic kidney disease, eye disease, dementia, depression, and all-cause mortality.

## Materials and Methods

### Study participants

UKB is a large-scale health research study with a long-term follow-up that began in 2006-2010 with the recruitment of approximately half a million adults in the UK. UKB was approved by the North West Multi-Center Research Ethics Committee. Written informed consent from all participants was obtained. At baseline, there were 405,319 middle-aged adults (aged 40-64 years), of whom 43,133 had prediabetes. Prediabetes was defined by a glycated hemoglobin (HbA1c) level of 5.7%-6.4% (39-47 mmol/mol) following the criteria of the ADA [17]. After exclusion of adults with prevalent cancer (N=2,386), and with missing data on frailty (N=127) and covariates (e.g., sex, educational level, etc; N=1,670), 38,950 middle-aged adults with prediabetes were included in the final analytic samples. Additionally, because the number of prevalent cases for each outcome varied, we assembled different analytic samples for each outcome (see details in **Figure 1**).

**Figure 1.**
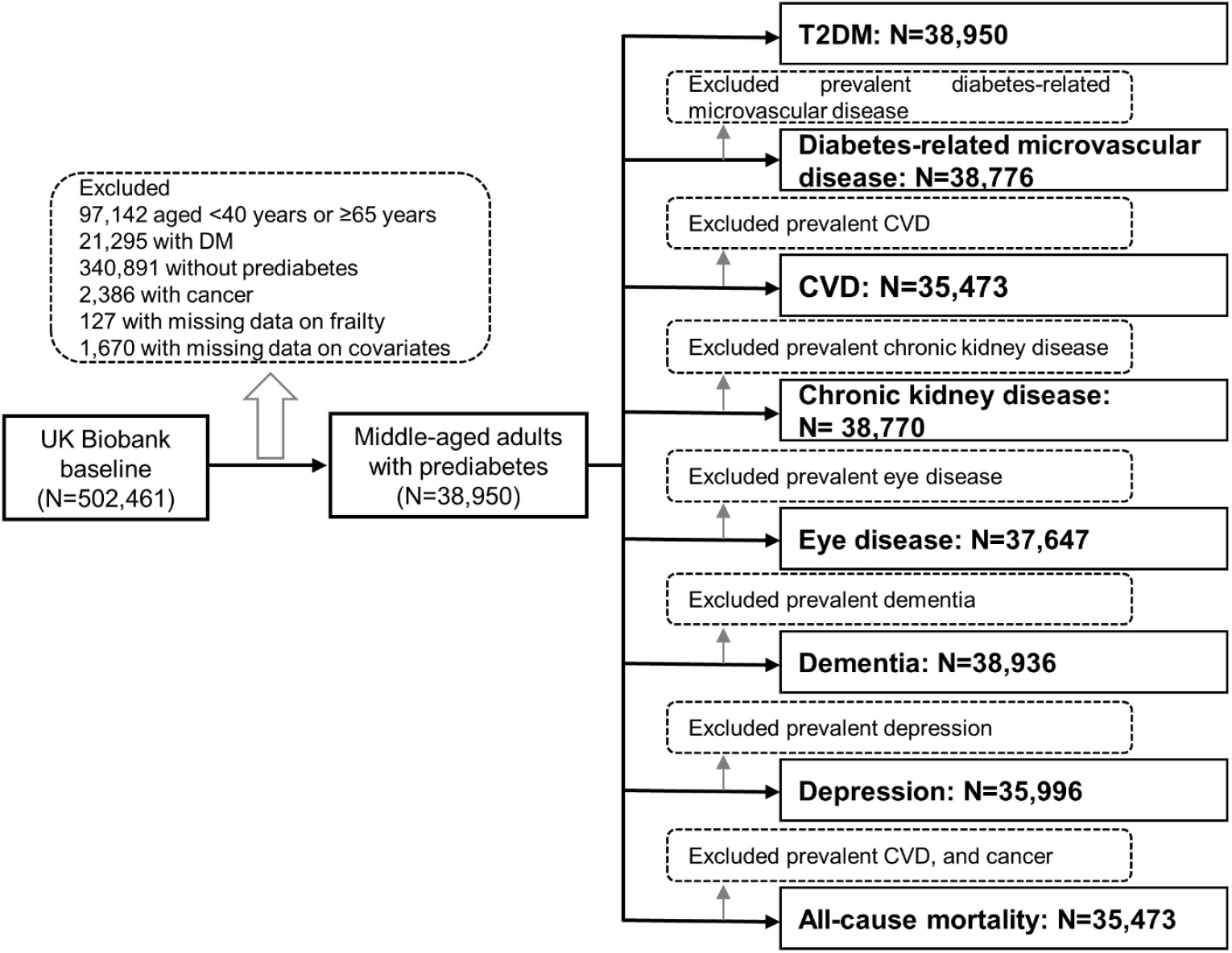
Flow chart of the analytic sample for analyses. Abbreviations: DM, diabetes mellitus; T2DM, type 2 diabetes mellitus; CVD, cardiovascular disease.

### Outcomes

In this study, the outcomes included T2DM, diabetes-related microvascular disease (contains retinopathy, neuropathy, and nephropathy), diabetes-related macrovascular disease (i.e., CVD, contains ischemic heart disease and stroke), chronic kidney disease, eye disease (contains cataract and glaucoma), mental disease (i.e., dementia, and depression), and all-cause mortality.

We defined T2DM using a UKB algorithm that combined self-reported medical history and medication information, as well as linked hospital admissions records (**Table S1**). In addition, according to the ADA criteria [17], undiagnosed T2DM cases were identified by random glucose (≥11.1 mmol/L) or HbA1c (≥6.5% [48 mmol/mol]). We ascertained incident cases of CVD, chronic kidney disease, eye disease, dementia, and depression by self-reported medical history (for the ascertainment of prevalent cases only) and linked hospital admissions records using the International Statistical Classification of Diseases and Related Health Problems 9^th^ (ICD-9) and 10^th^ (ICD-10) (**Table S1**). We ascertained death through linkage to national death registries. The time-to-event was calculated from the baseline (i.e., 2006-2010 years) to the occurrence of disease outcomes, death, loss to follow-up, or end of follow-up (2021 years), whichever came first.

### Frailty measurement

We used FP, a widely used physical frailty measurement proposed by Fried et al [8]. FP was evaluated by five criteria: unintentional weight loss, exhaustion, weakness, slow gait speed, and low physical activity, and has been previously applied in the UKB [18]. Of five criteria, weakness was assessed by objectively measured handgrip strength; the other four criteria were assessed by a self-reported questionnaire (see details in **Table 1**). The FP score ranged from 0 to 5, with a higher score indicating greater frailty. Participants were categorized into non-frail (FP score =0), pre-frail (1≤ FP score ≤2), and frail (FP score ≥3), as done in previous studies [8, 18].

**Table 1.**
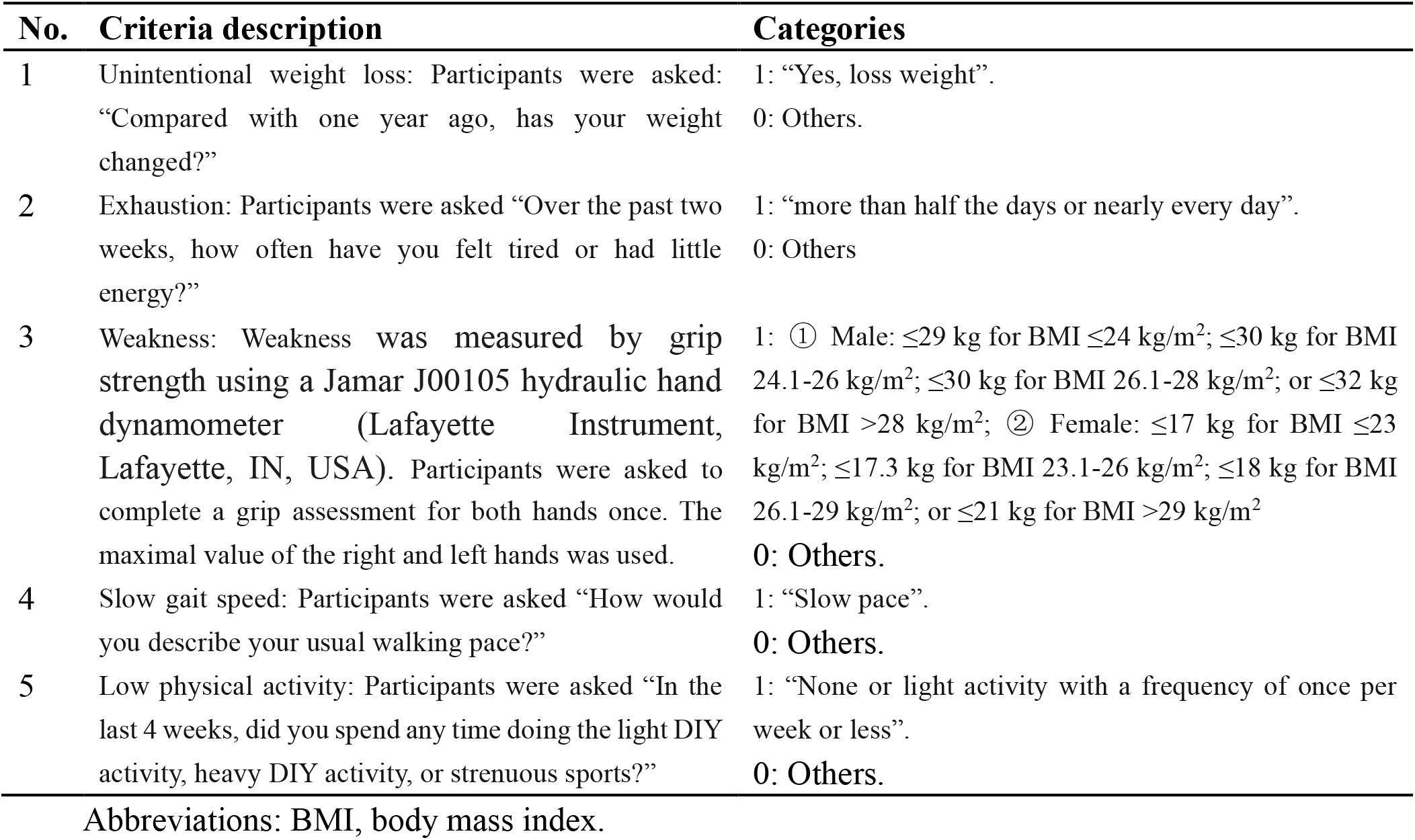
Five criteria for frailty phenotype in UK Biobank.

| **No.** | **Criteria description** | **Categories** |
| --- | --- | --- |
| 1 | Unintentional weight loss: Participants were asked: “Compared with one year ago, has your weight changed?” | 1: “Yes, loss weight”.<br>0: Others. |
| 2 | Exhaustion: Participants were asked “Over the past two weeks, how often have you felt tired or had little energy?” | 1: “more than half the days or nearly every day”.<br>0: Others |
| 3 | Weakness: Weakness was measured by grip strength using a Jamar J00105 hydraulic hand dynamometer (Lafayette Instrument, Lafayette, IN, USA). Participants were asked to complete a grip assessment for both hands once. The maximal value of the right and left hands was used. | 1: ① Male: $\leq 29$ kg for BMI $\leq 24$ kg/m<sup>2</sup>; $\leq 30$ kg for BMI 24.1-26 kg/m<sup>2</sup>; $\leq 30$ kg for BMI 26.1-28 kg/m<sup>2</sup>; or $\leq 32$ kg for BMI $> 28$ kg/m<sup>2</sup>; ② Female: $\leq 17$ kg for BMI $\leq 23$ kg/m<sup>2</sup>; $\leq 17.3$ kg for BMI 23.1-26 kg/m<sup>2</sup>; $\leq 18$ kg for BMI 26.1-29 kg/m<sup>2</sup>; or $\leq 21$ kg for BMI $> 29$ kg/m<sup>2</sup><br>0: Others. |
| 4 | Slow gait speed: Participants were asked “How would you describe your usual walking pace?” | 1: “Slow pace”.<br>0: Others. |
| 5 | Low physical activity: Participants were asked “In the last 4 weeks, did you spend any time doing the light DIY activity, heavy DIY activity, or strenuous sports?” | 1: “None or light activity with a frequency of once per week or less”.<br>0: Others. |

Abbreviations: BMI, body mass index.

### Covariates

Baseline data on age, sex (female, or male), ethnicity (Whites, Mixed, South Asian, Black, Chinese, or other background), educational level (high, intermediate, or low), occupational status (working, retired, or other), alcohol consumption (never or special occasions only, one to three times per month, one to four times per week, or daily or almost daily), smoking status (never, previous smoker, or current smoker), healthy diet (yes, or no), and family histories of diseases (including diabetes, CVD, dementia, and depression) were collected through a questionnaire interview. Townsend deprivation index (TDI) was calculated based on areas before participants were recruited in the UKB. Body mass index (BMI) was calculated as measured weight/height^2^ in kg/m^2^.

### Statistical analyses

Baseline characteristics of the analytic sample in total and by frailty status were presented as median (inter-quartile ranges [IQRs]) and number (percentage) for continuous variables and categorical variables, respectively. Kruskal-Wallis tests and Chi-square tests were used to compare the differences in characteristics by frailty status.

To evaluate the associations of frailty status (non-frail, pre-frail, and frail) with adverse outcomes, Cox proportional hazards regression models were performed. Schoenfeld residuals test was used to verify the proportional hazard assumption, and no significant violation was found. We calculated hazard ratios (HRs) and corresponding 95% confidence intervals (CIs) from two models. Model 1 was adjusted for age and sex. Model 2 was further adjusted for ethnicity, educational level, occupational status, TDI, alcohol consumption, smoking status, healthy diet, BMI, and family histories of diseases based on Model 1. Additionally, we calculated HRs (95% CIs) for adverse outcomes per 1-point increase in FP score.

Several sensitivity analyses were conducted to confirm the robustness of the results. First, to minimize the influence of reverse causality, we repeated the main analyses after excluding those without two years of follow-up. Second, to reduce the influence of poor health on frailty status, we repeated the main analyses after excluding participants with poor self-rated health status at baseline. Finally, we validated the associations of frailty with adverse outcomes among adults with T2DM. For adults with T2DM, HbA1c level (≥7.0% [≥53 mmol/mol], or <7.0% [<53 mmol/mol]), diabetes medication use (oral antidiabetes drug only, insulin, or neither), diabetes duration (in years), and prevalent diabetes-related microvascular disease (except for incident diabetes-related microvascular disease) were also included in Model 2.

We used SAS version 9.4 (SAS Institute, Cary, NC) to conduct all statistical analyses. To account for multiple testing, we used Bonferroni correction in all analyses (P<0.006).

## Results

### Baseline characteristics

Among 38,950 participants with prediabetes, the median age was 58.6 years (IQR: 53.1, 62.0), and the majority were female (54.3%) and White (89.1%) (**Table 2**). The prevalence of pre-frailty and frailty was 49.1%, and 5.9%, respectively (**Figure 2**). Pre-frail and frail adults were more likely to be female, have a lower educational level, and have a higher level of TDI and BMI, compared with the non-frail adults. **Table 2** shows the details of baseline characteristics by frailty status.

**Table 2.**
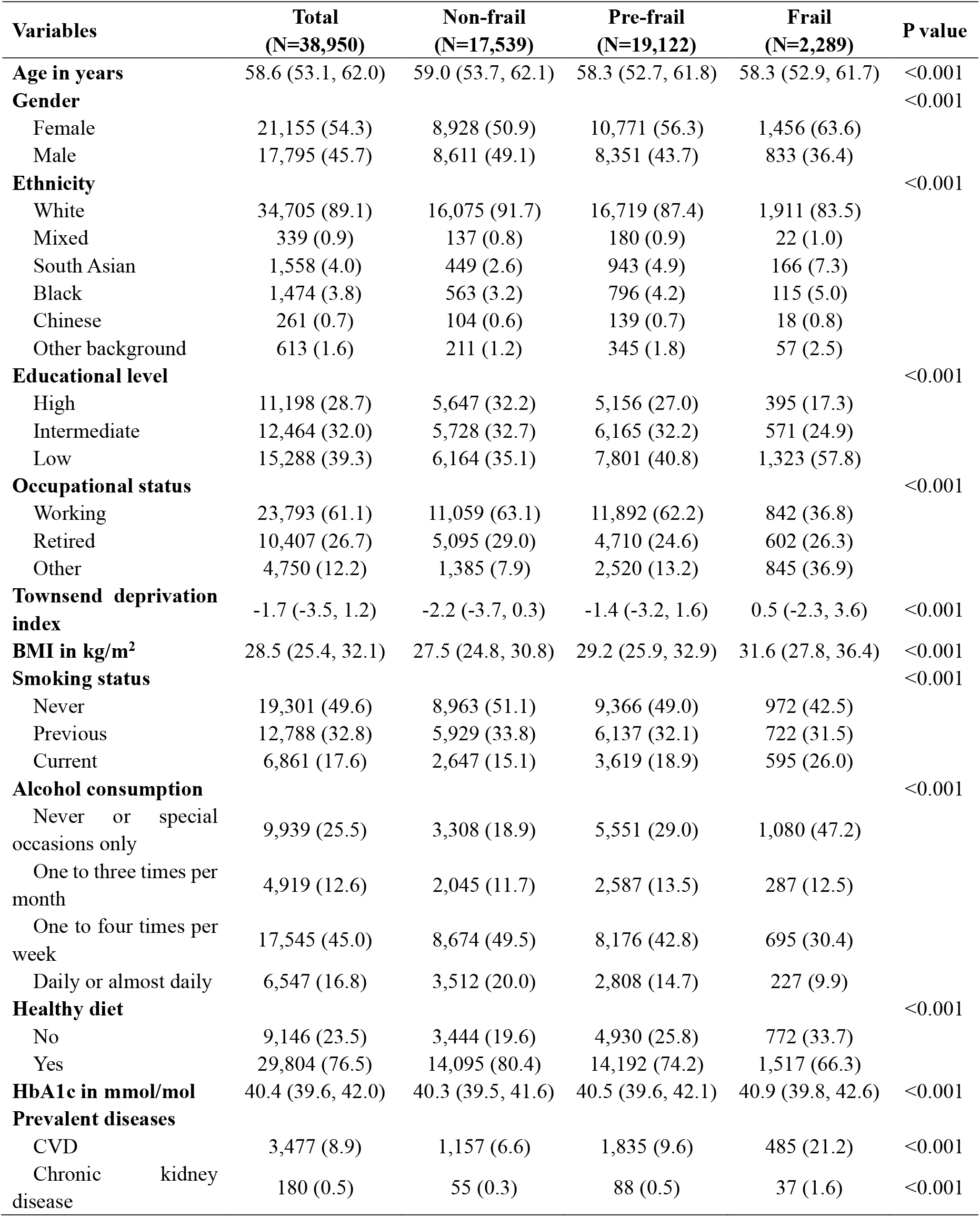
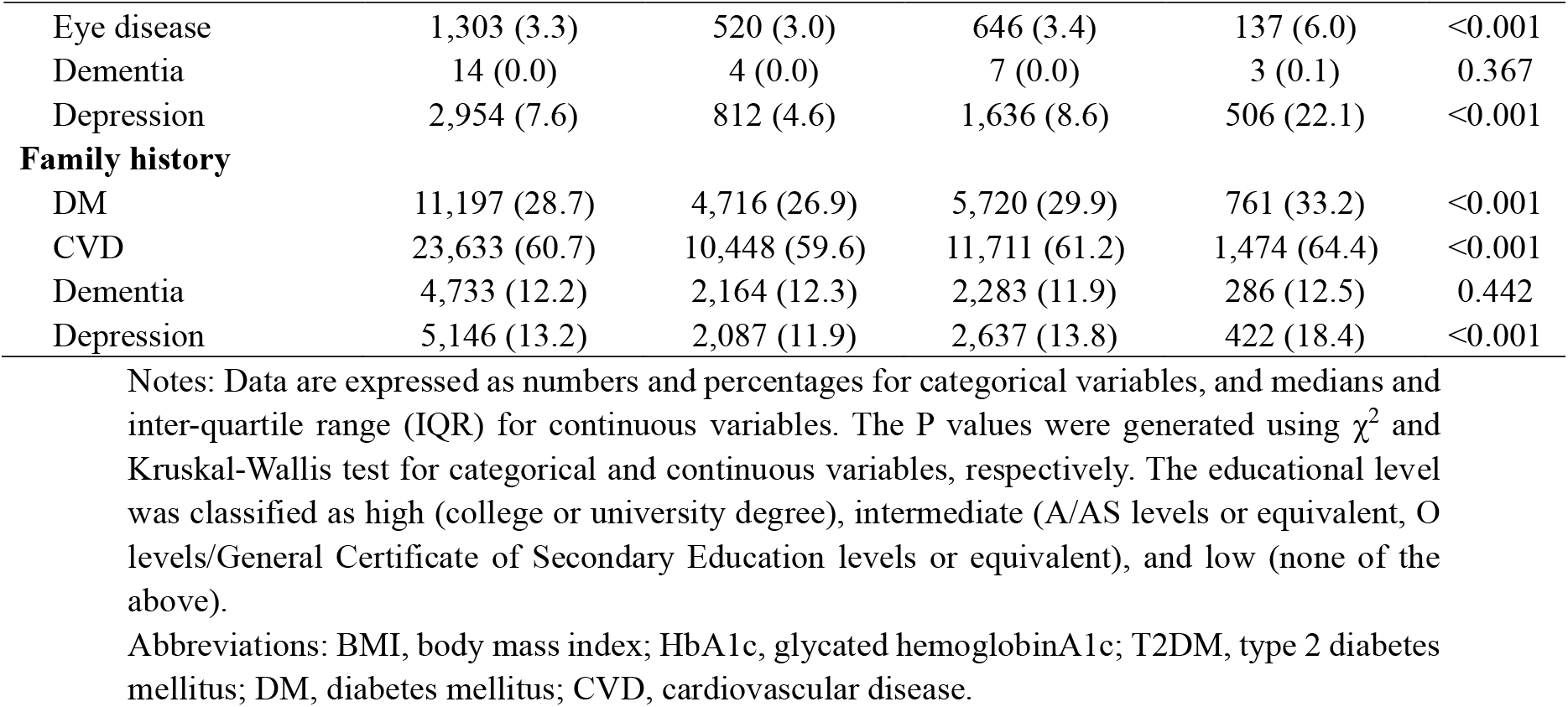
Baseline characteristics of study participants with prediabetes by frailty status.

**Figure 2.**
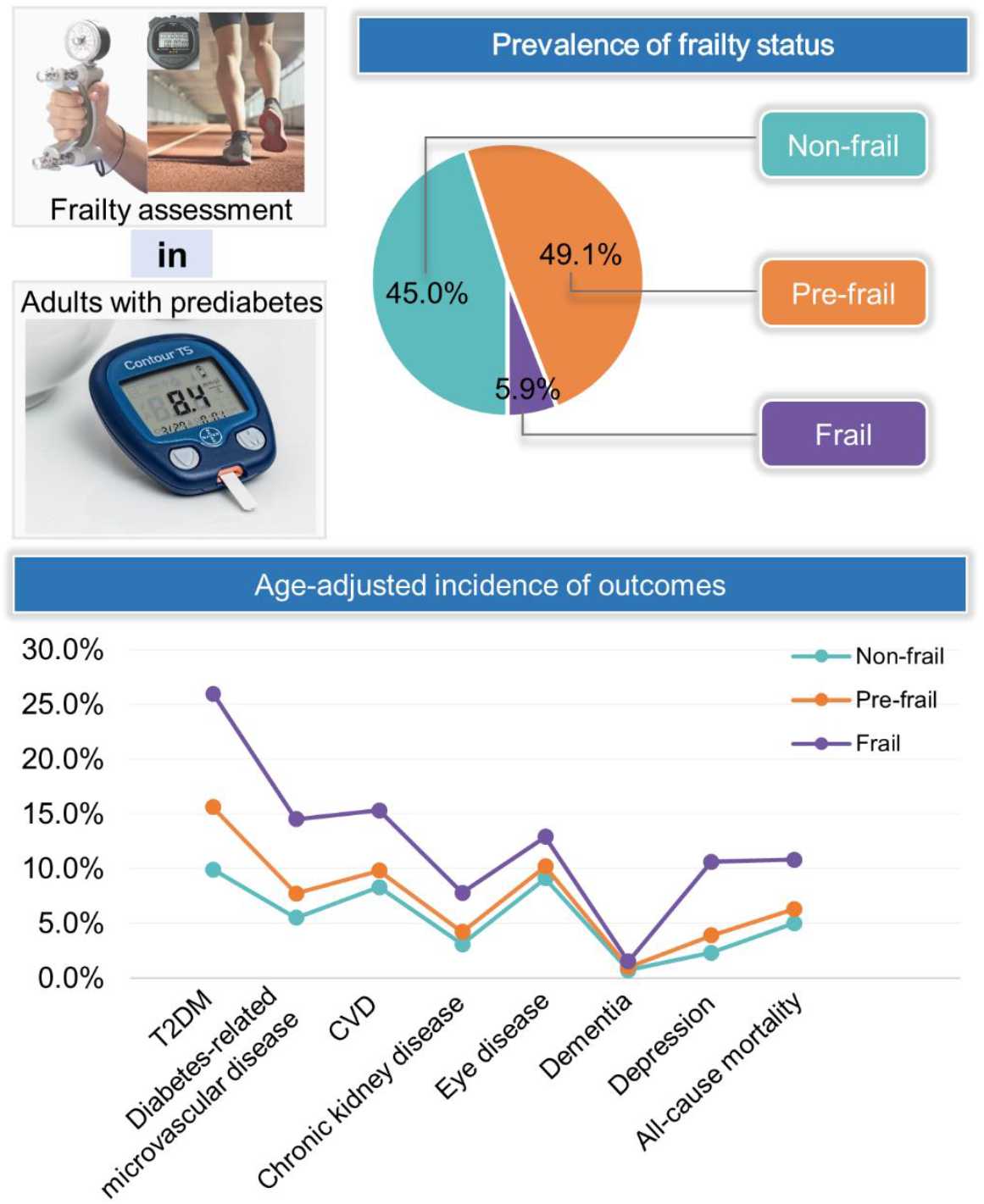
Prevalence of frailty status and age-adjusted incidence of adverse health outcomes among UKB participants with prediabetes during 12-years follow-up. Abbreviations: T2DM, type 2 diabetes mellitus; CVD, cardiovascular disease.

### Frailty and risks of adverse outcomes in middle-aged adults with prediabetes

During a mean follow-up of 446,662 persons-years, there were 5,289 incident T2DM, 2,657 incident diabetes-related microvascular disease, 3,234 incident CVD, 1,439 incident chronic kidney disease, 3,525 eye disease, 325 incident dementia, 1,265 incident depression, and 2,016 deaths. We found that frail participants developed more adverse outcomes than did the pre-frail and non-frail counterparts over the 12-year follow-up (**Figure 2**).

**Table 3** shows the associations of frailty with risks of multiple adverse outcomes in middle-aged adults with prediabetes. In the age- and sex-adjusted model, both pre-frailty and frailty were associated with higher risks of all adverse outcomes (all P for trend <0.001). After further adjusting for additional covariates, these associations remained statistically significant. When comparing pre-frail participants with their non-frail counterparts, the multivariable-adjusted HR was 1.35 (95% CI: 1.27, 1.43) for T2DM, 1.29 (95% CI: 1.18, 1.40) for diabetes-related microvascular disease, 1.17 (95% CI: 1.08, 1.26) for CVD, 1.22 (95% CI: 1.09, 1.37) for chronic kidney disease, 1.12 (95% CI: 1.04, 1.20) for eye disease, 1.57 (95% CI: 1.23, 2.01) for dementia, 1.48 (95% CI: 1.30, 1.68) for depression, and 1.25 (95% CI: 1.14, 1.38) for all-cause mortality. For frail participants, the multivariable-adjusted HR was 1.73 (95% CI: 1.55, 1.92) for T2DM, 1.89 (95% CI: 1.64, 2.18) for diabetes-related microvascular disease, 1.66 (95% CI: 1.44, 1.91) for CVD, 1.76 (95% CI: 1.45, 2.13) for chronic kidney disease, 1.31 (95% CI: 1.14, 1.51) for eye disease, 2.03 (95% CI: 1.33, 3.09) for dementia, 3.01 (95% CI: 2.47, 3.67) for depression, and 1.81 (95% CI: 1.51, 2.16) for all-cause mortality, compared to their non-frail counterparts. Additionally, with each 1-point increase in FP score, the incidence risks of these adverse outcomes significantly increased by 10%-42% (Model 2).

**Table 3.**
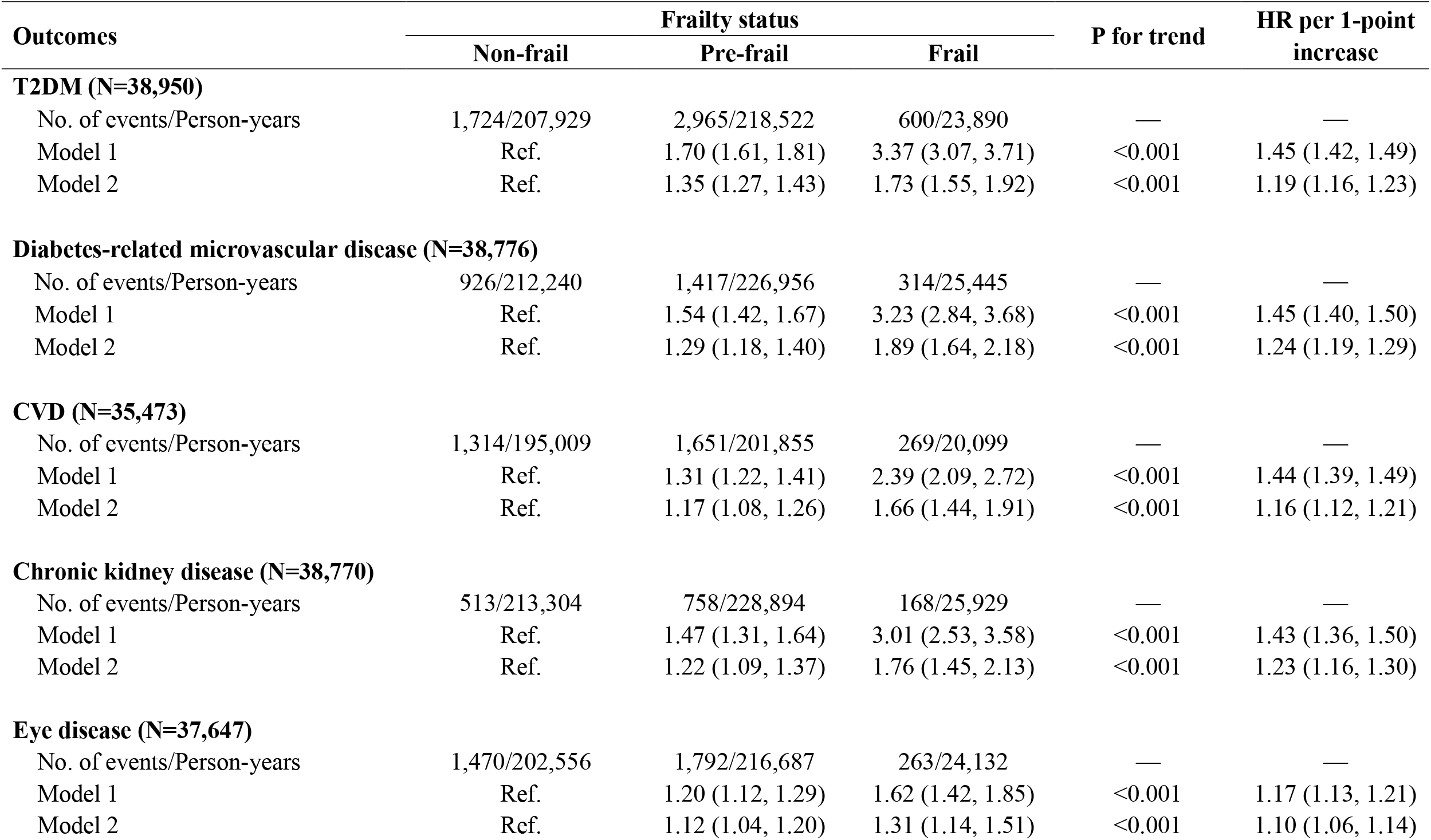
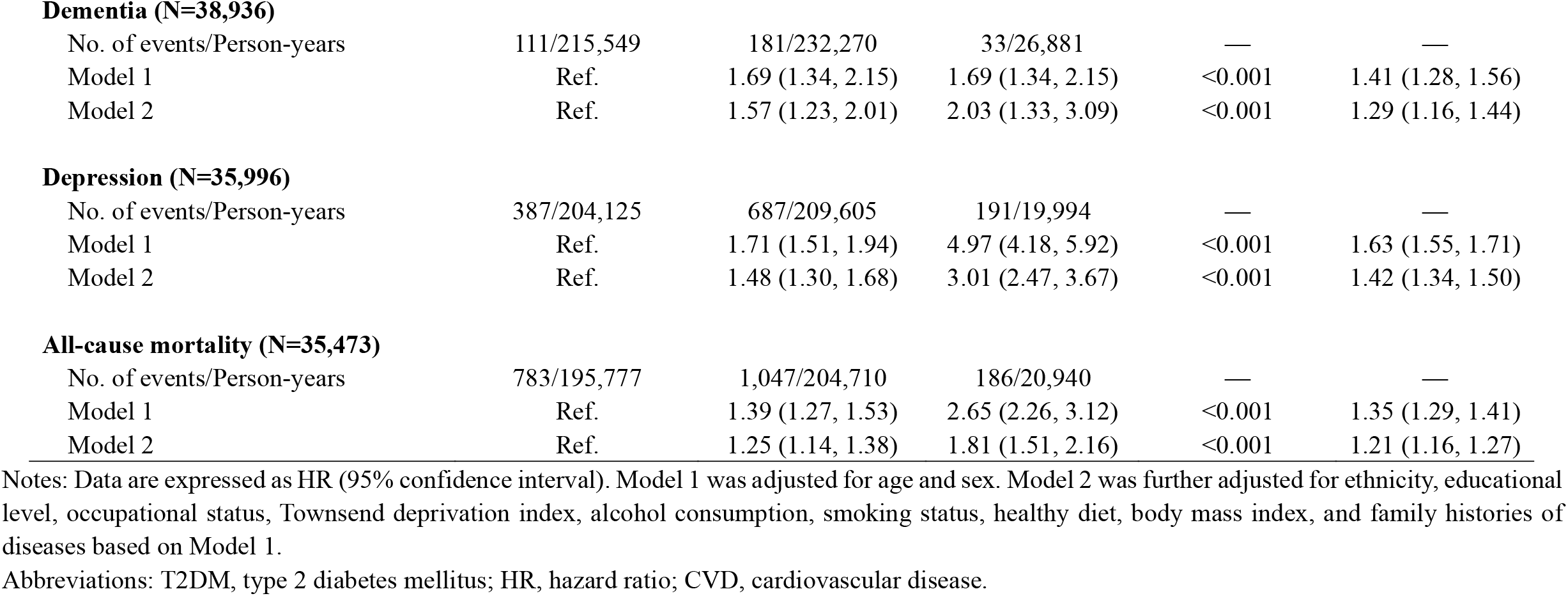
Associations of frailty with adverse health outcomes among middle-aged adults with prediabetes.

| Outcomes | Frailty status |  |  | P for trend | HR per 1-point increase |
| --- | --- | --- | --- | --- | --- |
|  | Non-frail | Pre-frail | Frail |  |  |
| **T2DM (N=38,950)** |  |  |  |  |  |
| No. of events/Person-years | 1,724/207,929 | 2,965/218,522 | 600/23,890 | — | — |
| Model 1 | Ref. | 1.70 (1.61, 1.81) | 3.37 (3.07, 3.71) | <0.001 | 1.45 (1.42, 1.49) |
| Model 2 | Ref. | 1.35 (1.27, 1.43) | 1.73 (1.55, 1.92) | <0.001 | 1.19 (1.16, 1.23) |
| **Diabetes-related microvascular disease (N=38,776)** |  |  |  |  |  |
| No. of events/Person-years | 926/212,240 | 1,417/226,956 | 314/25,445 | — | — |
| Model 1 | Ref. | 1.54 (1.42, 1.67) | 3.23 (2.84, 3.68) | <0.001 | 1.45 (1.40, 1.50) |
| Model 2 | Ref. | 1.29 (1.18, 1.40) | 1.89 (1.64, 2.18) | <0.001 | 1.24 (1.19, 1.29) |
| **CVD (N=35,473)** |  |  |  |  |  |
| No. of events/Person-years | 1,314/195,009 | 1,651/201,855 | 269/20,099 | — | — |
| Model 1 | Ref. | 1.31 (1.22, 1.41) | 2.39 (2.09, 2.72) | <0.001 | 1.44 (1.39, 1.49) |
| Model 2 | Ref. | 1.17 (1.08, 1.26) | 1.66 (1.44, 1.91) | <0.001 | 1.16 (1.12, 1.21) |
| **Chronic kidney disease (N=38,770)** |  |  |  |  |  |
| No. of events/Person-years | 513/213,304 | 758/228,894 | 168/25,929 | — | — |
| Model 1 | Ref. | 1.47 (1.31, 1.64) | 3.01 (2.53, 3.58) | <0.001 | 1.43 (1.36, 1.50) |
| Model 2 | Ref. | 1.22 (1.09, 1.37) | 1.76 (1.45, 2.13) | <0.001 | 1.23 (1.16, 1.30) |
| **Eye disease (N=37,647)** |  |  |  |  |  |
| No. of events/Person-years | 1,470/202,556 | 1,792/216,687 | 263/24,132 | — | — |
| Model 1 | Ref. | 1.20 (1.12, 1.29) | 1.62 (1.42, 1.85) | <0.001 | 1.17 (1.13, 1.21) |
| Model 2 | Ref. | 1.12 (1.04, 1.20) | 1.31 (1.14, 1.51) | <0.001 | 1.10 (1.06, 1.14) |
| **Dementia (N=38,936)** |  |  |  |  |  |
| No. of events/Person-years | 111/215,549 | 181/232,270 | 33/26,881 | — | — |
| Model 1 | Ref. | 1.69 (1.34, 2.15) | 1.69 (1.34, 2.15) | <0.001 | 1.41 (1.28, 1.56) |
| Model 2 | Ref. | 1.57 (1.23, 2.01) | 2.03 (1.33, 3.09) | <0.001 | 1.29 (1.16, 1.44) |
| **Depression (N=35,996)** |  |  |  |  |  |
| No. of events/Person-years | 387/204,125 | 687/209,605 | 191/19,994 | — | — |
| Model 1 | Ref. | 1.71 (1.51, 1.94) | 4.97 (4.18, 5.92) | <0.001 | 1.63 (1.55, 1.71) |
| Model 2 | Ref. | 1.48 (1.30, 1.68) | 3.01 (2.47, 3.67) | <0.001 | 1.42 (1.34, 1.50) |
| **All-cause mortality (N=35,473)** |  |  |  |  |  |
| No. of events/Person-years | 783/195,777 | 1,047/204,710 | 186/20,940 | — | — |
| Model 1 | Ref. | 1.39 (1.27, 1.53) | 2.65 (2.26, 3.12) | <0.001 | 1.35 (1.29, 1.41) |
| Model 2 | Ref. | 1.25 (1.14, 1.38) | 1.81 (1.51, 2.16) | <0.001 | 1.21 (1.16, 1.27) |

Notes: Data are expressed as HR (95% confidence interval). Model 1 was adjusted for age and sex. Model 2 was further adjusted for ethnicity, educational level, occupational status, Townsend deprivation index, alcohol consumption, smoking status, healthy diet, body mass index, and family histories of diseases based on Model 1.
Abbreviations: T2DM, type 2 diabetes mellitus; HR, hazard ratio; CVD, cardiovascular disease.

### Sensitivity analyses

Generally robust results were observed when excluding the participants with less than 2 years of follow-up (**Table S2**), or excluding the participants with poor self-rated health status at baseline (**Table S3**). In addition, we confirmed that frailty was positively associated with the risks of diabetes-related microvascular disease, CVD, chronic kidney disease, eye diseases, dementia, depression, and all-cause mortality in middle-aged adults with T2DM, and these associations were independent of factors related to diabetes severity at baseline (**Table S4**).

## Discussions

In a large sample of UKB participants with prediabetes, we, for the first time, demonstrated that both pre-frailty and frailty were associated with higher risks of multiple adverse outcomes, including T2DM, diabetes-related microvascular disease, CVD, chronic kidney disease, eye disease, dementia, depression, and all-cause mortality. Our findings support the heterogeneity of prediabetes in middle-aged adulthood, and suggest that assessing frailty status among middle-aged adults with prediabetes may help to identify those who were most at risk of subsequent adverse outcomes.

We observed a nearly twice higher prevalence of frailty in middle-aged adults with prediabetes (i.e., 5.9%) in comparison with general adults (i.e., 3.3%) [18]. Similarly, the prevalence of frailty among older adults with diabetes [19] is almost twice as high as that in those without diabetes (20.1% vs. 12%) [20]. It seems that adults with glucose metabolism disorders are experiencing accelerated aging process [21]. Multiple age-related metabolic disturbances are present in adults with prediabetes, including chronic inflammation, hyperglycemia, insulin resistance, and β-cell dysfunction [2, 15], creating a pathophysiological environment that contributes to frailty. Given the sharp increase of frailty after the age of 65 years [22], our findings suggest that there is a need for early identification of frailty, an aging indicator, in this middle-aged population with prediabetes.

To the best of our knowledge, this study was the first to report the associations of frailty with higher risks of a series of adverse outcomes in middle-aged adults with prediabetes. Only a few studies on the relationship between frailty and adverse outcomes included middle-aged adults with diabetes as a part of the study sample [23-25]. One prospective study of 998 African Americans aged 49-65 years has shown that frail diabetic adults had an increased risk of mortality [26]. The current large prospective study (N=38,950) showed that frailty was positively associated with higher risks of more outcomes including chronic kidney disease, eye disease, and dementia.

This study draws attention to the accelerated aging process in prediabetic adults, which may lead to rapid diabetes progression and contribute to the development of diabetes-related complications [21]. Nutritional and pharmacological anti-aging interventions have been revealed to help mitigate or reverse the accelerated aging process [27]. A recent review suggested that the most effective and easiest intervention strategy targeting frailty is to combine strength exercise and protein supplements in primary care [28]. Thus, our findings implicate that frailty assessment might help primary care providers identify the subpopulation at higher risk of adverse outcomes even in middle-aged prediabetic adults in communities. Next, early preventive and interventive programs targeting frailty in prediabetic adults are urgently needed. On the one hand, they may directly help reduce the occurrence of T2DM; on the other hand, may indirectly help reduce diabetes-related burdens. Meanwhile, pharmacologic intervention or other aggressive approaches to diabetes prevention are also encouraged [29, 30]. Before the formal implementation, much more research on the effectiveness and cost-effectiveness of interventional programs in this population is required.

The major strengths of this study were the large sample of middle-aged adults with prediabetes, long-term follow-up time, rich phenotype data, and linked hospital admissions records, enabling us to systematically evaluate the prospective associations of frailty with multiple adverse outcomes. There were several potential limitations. First, UKB was not representative of the sampling population, and the majority of included adults were Whites; thus, the results may not be generalizable to populations from other countries. Second, transitions in frailty status may occur over time [31], and evidence has suggested that transitions in frailty status were associated with adverse outcomes [32]. However, repeated measurements of frailty were lacking; thus, we were unable to estimate the influence of frailty transitions on the subsequent adverse outcomes in this study. Future longitudinal studies incorporating data on frailty transition are needed. Third, multiple outcomes were considered in this study, and thus, Type Ⅰ errors inevitably increased. To reduce the possibility of chance findings, we used Bonferroni correction. Finally, because of the observational study design, we could not draw a causal inference.

In this prospective cohort study of middle-aged adults with prediabetes, both pre-frailty and frailty were significantly associated with increased risks of multiple adverse outcomes, including T2DM, diabetes-related microvascular disease, CVD, chronic kidney disease, eye disease, mental disease, and all-cause mortality. The findings underscore the importance of frailty assessment in routine care for middle-aged adults with prediabetes. Detecting frailty at an early stage (i.e., accelerated aging) and implementing targeted interventions timely may help to improve the allocation of healthcare resources and to reduce diabetes-related burdens.

## Data Availability

Data from UK Biobank are available on application at www.ukbiobank.ac.uk/register-apply.

https://www.ukbiobank.ac.uk/enable-your-research/register

## Author contributions

ZL and Y. Zhu designed and coordinated the study. XC analyzed the data and drafted the article. XL, JZ, XS, GY, Y. Zhao, SL, EOH, XW, Y. Zhu, HA, TMG, and ZL contributed to the interpretation of data. XL, JZ, XS, GY, Y. Zhao, SL, EOH, XW, Y. Zhu, HA, TMG, and ZL revised the manuscript. All authors read and approved the final version of the manuscript.

## Data availability

Data from UK Biobank are available on application at www.ukbiobank.ac.uk/register-apply.

## Acknowledgments

This research has been conducted using the UK Biobank resource under application number 61856. We wish to acknowledge the UK Biobank participants who provided the sample that made the data available.

## References

1. IDF Diabetes Atlas. 10th edition. 2021; Available from: https://diabetesatlas.org/.

2. Tabak, A.G., et al., Prediabetes: a high-risk state for diabetes development. Lancet, 2012. 379(9833): p. 2279–90.

3. Schlesinger, S., et al., Prediabetes and risk of mortality, diabetes-related complications and comorbidities: umbrella review of meta-analyses of prospective studies. Diabetologia, 2022. 65(2): p. 275–285.

4. Harding, J.L., et al., Global trends in diabetes complications: a review of current evidence. Diabetologia, 2019. 62(1): p. 3–16.

5. Whicher, C.A., S. O’Neill, and R.I.G. Holt, Diabetes in the UK: 2019. Diabet Med, 2020. 37(2): p. 242–247.

6. Rooney, M.R., et al., Risk of Progression to Diabetes Among Older Adults With Prediabetes. JAMA Intern Med, 2021. 181(4): p. 511–519.

7. Lam, K. and S.J. Lee, Prediabetes-A Risk Factor Twice Removed. JAMA Intern Med, 2021. 181(4): p. 520–521.

8. Fried, L.P., et al., Frailty in older adults: evidence for a phenotype. J Gerontol A Biol Sci Med Sci, 2001. 56(3): p. M146–56.

9. Sergi, G., et al., Pre-frailty and risk of cardiovascular disease in elderly men and women: the Pro.V.A. study. J Am Coll Cardiol, 2015. 65(10): p. 976–83.

10. Soysal, P., et al., Relationship between depression and frailty in older adults: A systematic review and meta-analysis. Ageing Res Rev, 2017. 36: p. 78–87.

11. Aguayo, G.A., et al., Comparative analysis of the association between 35 frailty scores and cardiovascular events, cancer, and total mortality in an elderly general population in England: An observational study. PLoS Med, 2018. 15(3): p. e1002543.

12. Fan, J., et al., Frailty index and all-cause and cause-specific mortality in Chinese adults: a prospective cohort study. Lancet Public Health, 2020. 5(12): p. e650–e660.

13. Clegg, A. and Z. Hassan-Smith, Frailty and the endocrine system. Lancet Diabetes Endocrinol, 2018. 6(9): p. 743–752.

14. Abdul-Ghani, M.A., D. Tripathy, and R.A. DeFronzo, Contributions of beta-cell dysfunction and insulin resistance to the pathogenesis of impaired glucose tolerance and impaired fasting glucose. Diabetes Care, 2006. 29(5): p. 1130–9.

15. Lu, Q., et al., Community-based population data indicates the significant alterations of insulin resistance, chronic inflammation and urine ACR in IFG combined IGT group among prediabetic population. Diabetes Res Clin Pract, 2009. 84(3): p. 319–24.

16. Chhetri, J.K., et al., The prevalence and incidence of frailty in Pre-diabetic and diabetic community-dwelling older population: results from Beijing longitudinal study of aging II (BLSA-II). BMC Geriatr, 2017. 17(1): p. 47.

17. American Diabetes, A., 2. Classification and Diagnosis of Diabetes: Standards of Medical Care in Diabetes-2021. Diabetes Care, 2021. 44(Suppl 1): p. S15–S33.

18. Hanlon, P., et al., Frailty and pre-frailty in middle-aged and older adults and its association with multimorbidity and mortality: a prospective analysis of 493 737 UK Biobank participants. Lancet Public Health, 2018. 3(7): p. e323–e332.

19. Kong, L.N., et al., The prevalence of frailty among community-dwelling older adults with diabetes: A meta-analysis. Int J Nurs Stud, 2021. 119: p. 103952.

20. O’Caoimh, R., et al., Prevalence of frailty in 62 countries across the world: a systematic review and meta-analysis of population-level studies. Age Ageing, 2021. 50(1): p. 96–104.

21. Palmer, A.K., et al., Cellular senescence: at the nexus between ageing and diabetes. Diabetologia, 2019. 62(10): p. 1835–1841.

22. Raymond, E., et al., Drivers of Frailty from Adulthood into Old Age: Results from a 27-Year Longitudinal Population-Based Study in Sweden. J Gerontol A Biol Sci Med Sci, 2020. 75(10): p. 1943–1950.

23. Pandey, A., et al., Association of Baseline and Longitudinal Changes in Frailty Burden and Risk of Heart Failure in Type 2 Diabetes - Findings from the Look AHEAD Trial. J Gerontol A Biol Sci Med Sci, 2022.

24. Liccini, A. and T.K. Malmstrom, Frailty and Sarcopenia as Predictors of Adverse Health Outcomes in Persons With Diabetes Mellitus. J Am Med Dir Assoc, 2016. 17(9): p. 846–51.

25. Chao, C.T., et al., Both pre-frailty and frailty increase healthcare utilization and adverse health outcomes in patients with type 2 diabetes mellitus. Cardiovasc Diabetol, 2018. 17(1): p. 130.

26. Chode, S., et al., Frailty, Diabetes, and Mortality in Middle-Aged African Americans. J Nutr Health Aging, 2016. 20(8): p. 854–859.

27. Ros, M. and J.M. Carrascosa, Current nutritional and pharmacological anti-aging interventions. Biochim Biophys Acta Mol Basis Dis, 2020. 1866(3): p. 165612.

28. Travers, J., et al., Delaying and reversing frailty: a systematic review of primary care interventions. Br J Gen Pract, 2019. 69(678): p. e61–e69.

29. Diabetes Prevention Program Research, G., et al., 10-year follow-up of diabetes incidence and weight loss in the Diabetes Prevention Program Outcomes Study. Lancet, 2009. 374(9702): p. 1677–86.

30. Tuomilehto, J., et al., Prevention of type 2 diabetes mellitus by changes in lifestyle among subjects with impaired glucose tolerance. N Engl J Med, 2001. 344(18): p. 1343–50.

31. Anker, D., et al., How blood pressure predicts frailty transitions in older adults in a population-based cohort study: a multi-state transition model. Int J Epidemiol, 2021.

32. Wang, M.C., et al., Frailty, transition in frailty status and all-cause mortality in older adults of a Taichung community-based population. BMC Geriatr, 2019. 19(1): p. 26.

